# Estimating the Effects of Treatment Regimes over the Course of Chronic Disease: A Multi-state Causal Framework with Baseline Confounding

**DOI:** 10.1101/2025.07.25.25332203

**Authors:** Ming Ding

## Abstract

The development of chronic disease is a long-term process that involves multiple endpoints, and few methods can assess the health benefits of a treatment regime over the disease course. Existing multi-state Cox models estimate survival risks by state over time, which are difficult to use when comparing the effectiveness of treatment regimes. A discrete-time split-state framework has been proposed, which divides disease states into substates by conditioning on past history. As this framework is both “memoryless” and “memorable”, the time-specific transition parameters can be synthesized into summary measures, substate-specific life year (SSLY), multimorbidity-adjusted life year (MALY), and disease path. In this paper, based on this framework, we propose to investigate the causal effects of static and dynamic treatment regimes on health benefits over the entire disease course, under the assumptions of constant confounders from baseline and instantaneous effects of interventions on transition rates. Our method can identify the optimal treatment regime that generates the most benefits using MALY, and illustrate the mechanisms of treatment regimes affecting disease progression using SSLY and disease path. In the application, we evaluated the cardiovascular benefits of smoking cessation using data from the Atherosclerosis Risk in Communities (ARIC) study, where the course of heart disease was modeled in healthy (S_0_), at metabolic risk (S_1_), coronary heart disease (S_2_), heart failure (S_3_), and mortality states (S_4_). Compared to the regime “being a smoker in S_0_-S_4_”, the MALY was 0.53 (95% CI: 0.21, 0.96), 6.10 (95% CI: 4.88, 7.19), and 4.34 (95% CI: 3.02, 5.47) years higher for the regimes “being a smoker in S_0_ and S_1_ and stop smoking if a person develops S_2_, S_3,_ or S_4_”, “no smoking in S_0_-S_4_”, and “being a smoker at the start of intervention and stop smoking if age>65y”, respectively. In summary, our method can evaluate the health benefits of treatment regimes over the disease course, and has the potential to improve the precision of chronic disease prevention.

## 1. INTRODUCTION

Comparative effectiveness of interventions has been widely studied in medical research. The interventions include static regimes, where all treatment during follow up are predetermined at baseline, and dynamic regimes, where the treatment at each time point depends on the individual’s response to previous treatments. The health effect of a treatment regime is measured by the risk of disease development using the parametric g-formula or the marginal structure model (MSM).^1,2^ These models first estimate conditional survival risk within each stratum of confounders, and then obtain the marginal survival risk for the entire population using standardization (g-formula) or inverse probability weighting (MSM).

Rather than being based on a single disease endpoint, the development of chronic disease is a long-term process that involves multiple endpoints. Taking heart disease as an example, it begins with the development of hypertension, hypercholesterolemia, or hyperglycemia (i.e., an at-metabolic-risk state), followed by the onset of coronary heart disease (CHD), which may progress to heart failure and eventually end in mortality.^3,4^ The disease states in this multi-state process are interconnected, and treatment at each state can influence the entire course of disease progression. However, existing methods that assess the effects of treatment regimes based on a single endpoint overlook the disease course and may misidentify the optimal treatment.

Multi-state modeling has been used to describe the course of chronic diseases. For example, multi-state Cox models apply multiple Cox models to describe state transitions;^5^ however, the estimated transition parameters are state- and time-specific, making it difficult to use them to evaluate the effects of treatment regimes. A discrete-time split-state framework for multi-state modeling has been proposed,^6^ which divides disease states into substates by conditioning on past history. As the substates contain information about past history, the time-specific transition parameters can be synthesized into summary measures, multimorbidity-adjusted life year (MALY), substate-specific life year (SSLY), and disease path.^7^ In this paper, based on this framework, we develop a multi-state g-formula to evaluate the health benefits of treatment regimes over the disease course. Our method can identify the optimal treatment regime that generates the most benefits as indicated by MALY, and illustrate the mechanisms of treatment regimes on disease progression as indicated by SSLY and disease path. Our method is developed under the assumptions of constant confounders from baseline and instantaneous effects of interventions on transition rates.

## 2. METHODS

### 2.1 An introduction to the Discrete-Time Multi-State Framework

We apply a Discrete-Time Multi-State Framework to estimate the health benefits of treatment regime over the disease course.^6,7^ While it can be used for chronic disease of any states, we illustrate it using five states S_0_-S_4_ (**Fig 1**). In brief, the framework divides disease state into substates by conditioning on past history and estimates transition rates between substates using cause-specific Cox models. For example, S_2_ can be divided into substates S_2|0_ and S_2|1_0_, where S_2|0_ is transitioned from S_0_ and S_2|1_0_ is transitioned from S_0_ and S_1_. The framework has two unique features. First, by conditioning on past states, the newly created substates are independent of past history and exhibit the Markov properties. Thus, regardless of whether the original process is Markov, the Aalen-Johansen estimator can be used to estimate state occupation probabilities based on transition rates. Second, while satisfying the Markov assumption, the substates contain information on past history and indicate multimorbidity status. Thus, the time-specific transition rates and state occupation probabilities can be synthesized into summary measures, SSLY, MALY, and disease path.**^7^**

**Figure 1.**
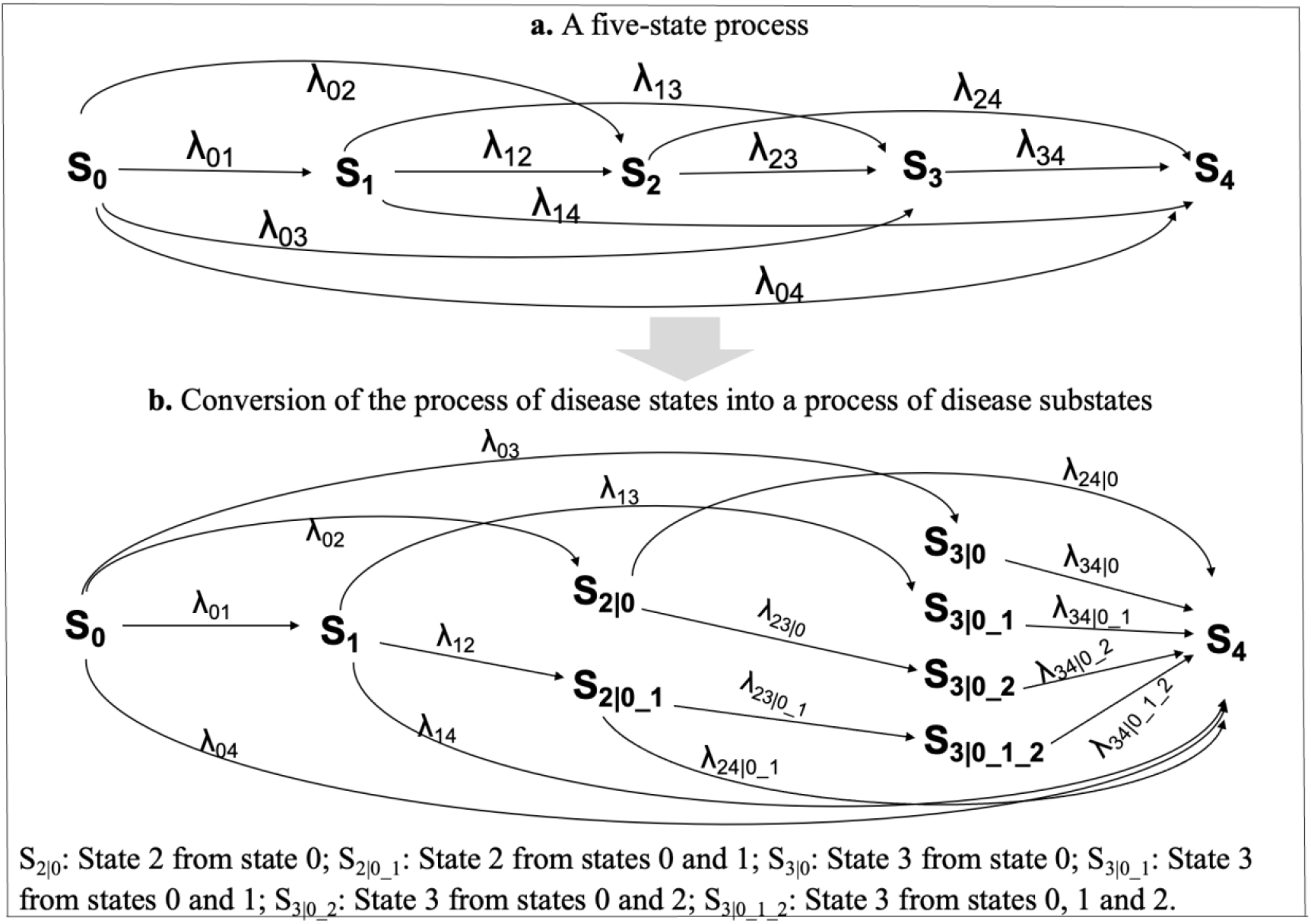
Construction of the process of disease substates by conditioning on past states. Note: This figure is cited from a previous publication.[1] [1] Ding M., Chen H., F.C. L. A discrete-time split-state framework for multi-state modeling with application to describing the course of heart disease. BMC Medical Research Methodology. 2025:25-54.

**Figure 2.**
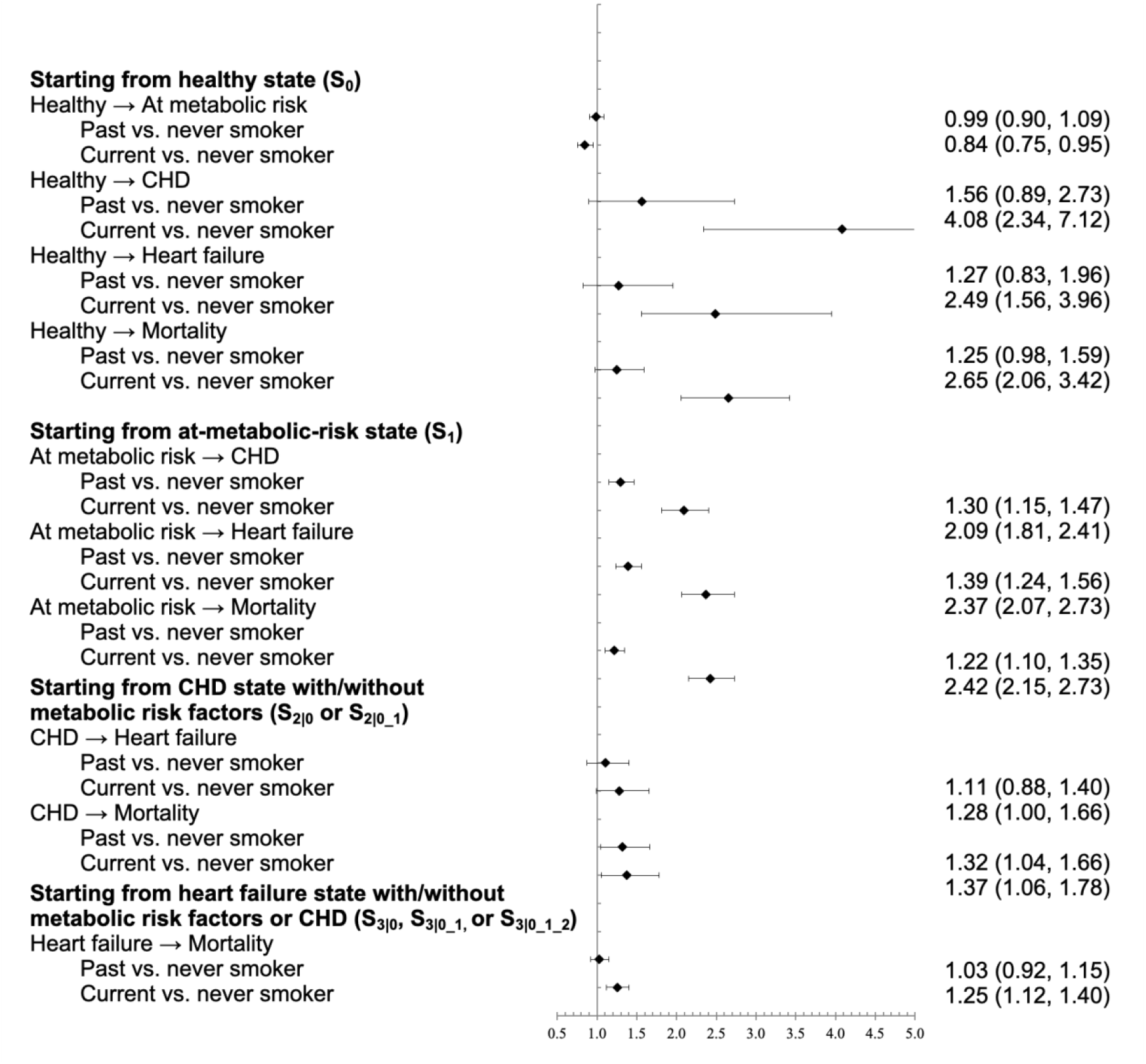
Hazard ratios (95% CIs) for transition by smoking status in each disease substate in the Atherosclerosis Risk in Communities (ARIC) Study starting from 1987 to 2019 (n=15027).

Suppose we have a population of N participants. Let *m* denote disease states, and a disease process includes states *S*_0_, *S*_1_, *S*_*m*_, …, and *S*_*M*_. By conditioning on past history, the five states *S*_0_, *S*_1_, *S*_2_, *S*_3_, and *S*_4_ can be divided into nine disease substates, *S*_0_, *S*_1_, *S*_2|0_, *S*_2|0_1_, *S*_3|0_, *S*_3|0_1_, *S*_3|0_2_, *S*_3|0_1_2_, and *S*_4_. Let *E*_0_, *E*_1_, …, *E*_*M*−1_ refer to exposure in states *S*_0_, *S*_1_, *S*_*m*_, …, and *S*_*M*−1_ at time *t* (Note: there is no exposure in the last state, *S*_*M*_). By dividing into substates, the exposures are denoted as *E*_0_, *E*_1_, *E*_2|0_, *E*_2|0_1_, *E*_3|0_, *E*_3|0_1_, *E*_3|0_2_, and *E*_3|0_1_2_. Let ***L*** be confounders at baseline. Our method is developed under two assumptions: 1) ***L*** are constant over the follow up period; 2) The effects of the exposure on the transition rates of developing the next states are instantaneous.

### 2.2 Estimate conditional transition rates

As described in detail in previous publication, cause-specific Cox models are applied to examine the associations between exposure and transition rates. In brief, the data are divided into subsets by disease state: Subset 1 starts from S_0_=1 and ends in the incidence of S_1_, S_2,_ S_3_ or S_4_; subset 2 starts from S_1_=1 and ends in the incidence of S_2,_ S_3_ or S_4_; subset 3 starts from S_2_=1 and ends in the incidence of S_3_ or S_4_; and subset 4 starts from S_3_=1 and ends in the incidence of S_4_ or the end of follow up, whichever comes first. Each data subset is restructured from a wide to long format by age and cause-specific Cox models (**Models 1-4**) are fitted to each data subset to estimate conditional transition rates.

**Model 1** in subset 1 starting from S_0_ state:

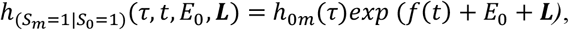

where *m* ranges from 1-4, indicating the development of S_1_, S_2_, S_3_, or S_4_ states from S_0_. *f*(*t*) is a function of age. *h*_0*m*_ (*τ*) is the baseline hazard of developing state *m. τ* is time to event in each time interval and ranges from [0, 1] (we use age as the time scale, and thus the maximum length of each time interval is 1).

**Model 2** in subset 2 starting from S_1_ state:

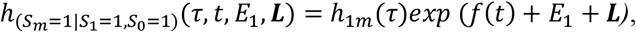

where *m* can be 2-4, indicating the development of S_2_, S_3_, or S_4_ states.

**Model 3** in subset 3 starting from S_2_ state:

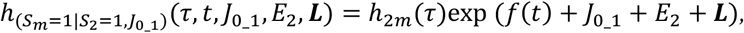

where *m* can be 3 and 4, indicating the development of S_3_ or S_4_ states from S_2_. J_0_1_ is a joint indicator of past states S_0_ and S_1_, and has two categories, “0” and “0_1”. For J_0_1_ = “0”, *E*_2_ equals to *E*_2|0_, and for J_0_1_ = “0_1”, *E*_2_ equals to *E*_2|0_1_.

**Model 4** in subset 4 starting from S_3_ state:

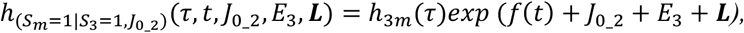

where *m* indicates the development of S_4_ state from S_3_. J_0_2_ is a joint indicator of past states S_0_, S_1_ and S_2_, and has 4 categories, “0”, “0_1”, “0_2”, “0_1_2”. For J_0_2_=“0”, “0_1”, “0_2”, “0_1_2”, *E*_3_ refers to *E*_3|0_, *E*_3|0_1_, *E*_3|0_2_, and *E*_3|0_1_2_, respectively.

The discrete-time hazard can be approximated by age-specific risk at the end of each time interval,^8^ which can be estimated using the cumulative incidence function (CIF) of cause-specific Cox models.^9,10^ Specifically, **Model 1** estimates *λ*_01_(*t*|*E*_0_, ***L****), λ*_02_(*t*|*E*_0_, ***L****), λ*_03_(*t*|*E*_0_, ***L***), and *λ*_04_(*t*|*E*_0_, ***L***), conditional transition rates from S_0_ to states S_1_ - S_4_, respectively. **Model 2** estimates *λ*_12_(*t*|*E*_1_, ***L****), λ*_13_(*t*|*E*_1_, ***L***), and *λ*_14_(*t*|*E*_1_, ***L***), conditional transition rates from S_1_ to states S_2_ - S_4_, respectively. **Model 3** estimates *λ*_23|0_(*t*|*E*_2|0_, ***L***) and *λ*_24|0_(*t*|*E*_2|0_, ***L***), conditional transition rates from S_2|0_ to S_3_ and S_4_, respectively, and *λ*_23|0_1_(*t*|*E*_2|0_1_, ***L***) and *λ*_24|0_1_(*t*|*E*_2|0_1_, ***L***), conditional transition rates from S_2|0_1_ to S_3_ and S_4_, respectively. **Model 4** estimates *λ*_34|0_(*t*|*E*_3|0_, ***L****), λ*_34|0_1_(*t*|*E*_3|0_1_, ***L****), λ*_34|0_2_(*t*|*E*_3|0_2_, ***L***), and *λ*_34|0_1_2_(*t*|*E*_3|0_1_2_, ***L***), conditional transition rates to S_4_ from S_3|0_, S_3|0_1_, S_3|0_2_, and S_3|0_1_2_, respectively. These conditional transition rates can be used to construct the conditional transition rate matrix ***Q***(*t*|***E***, ***L)***as shown below, where ***E*** = (*E*_0_, *E*_1_, *E*_2|0_, *E*_2|0_1_, *E*_3|0_, *E*_3|0_1_, *E*_3|0_2_, *E*_3|0_1_2_). Each item of the matrix shows the average transition rates from the substate in the row to the substate in the column in a population.

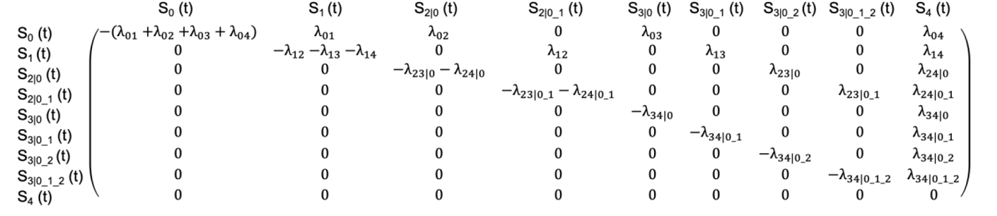

### 2.3. Static treatment regime and conditional SSLY, MALY, and disease path

Let ***A*** denote treatment regime with ***A*** = (*A*_0_, *A*_1_, *A*_2|0_, *A*_2|0_1_, *A*_3|0_, *A*_3|0_1_, *A*_3|0_2_, *A*_3|0_1_2_). For static regime, the hypothetical intervention A is constant over time. Let ***P***(t|***A*, L)** denote conditional state occupation probabilities at time t. For a five-state transition, ***P***(t|***A*, L)** = c(p_0_(t), p_1_(t), p_2|0_(t), p_2|1_0_(t), p_3|0_(t), p_3|1_0_(t), p_3|2_0_(t), p_3|2_1_0_(t), p_4_(t)), which are the percentages of participants occupied in *S*_0_, *S*_1_, *S*_2|0_, *S*_2|0_1_, *S*_3|0_, *S*_3|0_1_, *S*_3|0_2_, *S*_3|0_1_2_, and *S*_4_ substates at time *t*. By using the Aalen-Johansen estimator,^11^ conditional state occupation probability ***P***(t|***A*, L**) can be estimated as 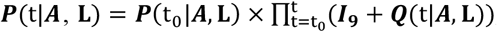, where ***I*_9_**is an identity matrix, and t_0_ is the start of follow up.

The estimation of SSLY and MALY based on ***P***(t|***A*, L**) has been shown in detail in previous publication.^7^ In brief, the substate-specific life year (SSLY) can be estimated as the sum of probabilities of being in each substate (except S_4_) from the starting age until death. Let *SSLY*(***A*, L**) be a 1× *k* matrix denoting life year in *k* substates, where *k* ranges from 1-8, referring to substates *S*_0_, *S*_1_, *S*_2|0_, *S*_2|0_1_, *S*_3|0_, *S*_3|0_1_, *S*_3|0_2_, and *S*_3|0_1_2_, respectively. The SSLY in substate *k* is estimated as 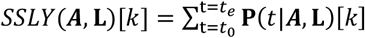, where *t*_0_ is the start of follow up and *t*_*e*_is the end.

As substates indicate multimorbidity status, a disability weight (DW) is assigned to each substate to account for the severity of multimorbidity.^12,13^ As all substates are mutually exclusive, the conditional MALY can be calculated as a weighted average of SSLY across *k* substates.

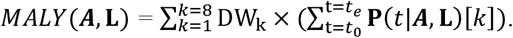

Let *PATH*(***A*, L**) be a 1× *K* matrix denoting death from the *K* substates. The estimation of disease path has also been shown in detail in previous publication.^7^ In brief, the increase in probability in the death state, Δp_4_(t), is estimated as

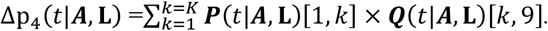

As ***P***(*t*|***A*, L**)[1, *k*] × ***Q***(*t*|***A*, L**)[*k*, 9] traces the increase in the probability of death from substate *k* at age t. The increase in the probabilities of death from substate *k* starting from age *t*_1_ to *t*_*e*_can be estimated as

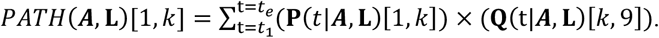

For a five-state process, the 1 to 8^th^ columns of *PATH*(***A*, L**) indicate the probabilities of death transited from substates S_0_, S_1_, S_2|0_, S_2|0_1_, S_3|0_, S_3|0_1_, S_3|0_2_, and S_3|0_1_2_, respectively. As disease substates track past history, these eight columns of *PATH*(***A*, L**) indicate the probabilities of death from paths S_0_→S_4_, S_0_→S_1_→S_4_, S_0_→S_2_→S_4_, S_0_→S_1_→S_2_→S_4,_ S_0_→S_3_→S_4_, S_0_→S_1_→S_3_→S_4_, S_0_→S_2_→S_3_→S_4_, and S_0_→S_1_→S_2_→S_3_→S_4_, respectively.

### 2.4 Static treatment regime and counterfactual SSLY, MALY, and disease path

Theoretically, the counterfactual MALY of a treatment regime is the average MALY in a population had everyone received the treatment regime.^8^ This is essentially to estimate the standardized MALY according to the distribution of **L** in a population. The population-level counterfactual MALY can be estimated as *MALY***^*A*^** = ∫ *MALY*(***A, L***)*f*(***L***)*d****L***, where *f*(**L**) is the joint density of confounders **L**. -**Formula 1** However, in practice, it is difficult to estimate *f*(**L**) if **L** are continuous variables. In fact, **Formula 1** is equivalent to the individual-level estimation of counterfactual MALY, with the illustration and proof shown below.

We estimate the conditional transition rates for each person from **Models 1-4**. For person *i*, as we only observe the person being in one substate *s*_*m*(*i*)_(*t*) with exposure *e*_*m*(*i*)_(*t*) at time *t*, we can estimate conditional transition rates as *λ*_***m***(***i***)_(*t*|***e***_***m***(***i***)_(*t*), ***L*_*i*_**), and construct the counterfactual transition rate matrix 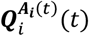 under the hypothetical intervention.

The counterfactual transition rates are used to construct the matrix 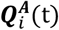, and the counterfactual state occupation probability is estimated as

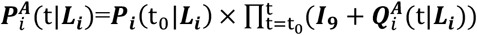

For person *i*, the counterfactual SSLY in substate *k* is estimated as

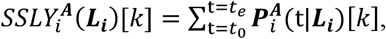

For person *i*, the counterfactual MALY is estimated as

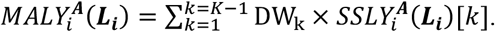

For person *i*, the counterfactual disease path in substate *k* is estimated as

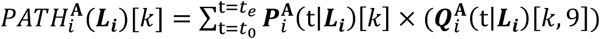

The counterfactual SSLY in substate *k* in the whole population can be estimated as

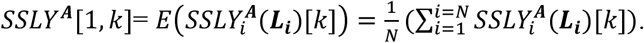

The counterfactual MALY in the whole population can be estimated as

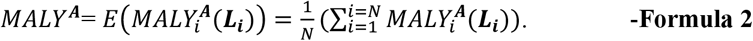

In fact, the population-level estimation (**Formula 1)** and individual-level estimation (**Formula 2)** of counterfactual MALY are equivalent, with proof shown below.

**Proof.** We divide **L** into *P* stratum ***l*_1_, *l*_2_**, …, ***l***_***P***_, with *n*_1_, *n*_2_, …, *n*_*P*_participants in each stratum.

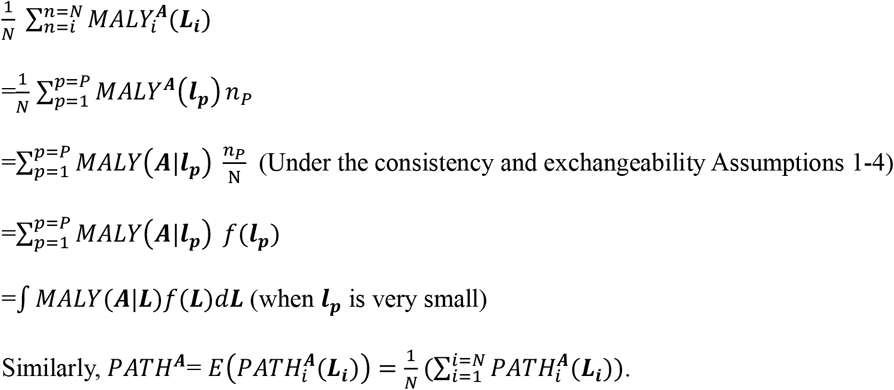

### 2.5. Dynamic treatment regime and individual-level estimation of counterfactual MALY

The dynamic treatment regime ***A***_*i*(*t*)_can vary over time by person, depending on the decision criteria, such as side effects of treatment. Let *W*_*i*(*t*)_indicate the side effects at time *t*, which can be affected by ***A***_*i*(*t*)_and baseline confounders ***L***_*i*_. We assume that intervention ***A***_*i*(*t*)_only has instantaneous effect on *W*_*i*(*t*)_(i.e., the effect is only at *t* and not the time beyond).

**Step 1**. For person *i* starting from *t*_0_, based on the state occupation probability ***P*_*i*_**(*t*_0_|***L*_*i*_**), we assign treatment ***A***_*i*(*t*0)_.

**Step 2**. The treatment ***A***_*i*(*t*0)_is used to fit **Models 1**−**4** and construct counterfactual transition rate matrix 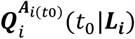.

**Step 3.** The state occupation probabilities for participant *i* at *t*_1_ is estimated as

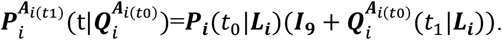

**Step 4**. The adverse effect at *t*_1_, *W*_*i*(*t*1)_, can be estimated as *E*(*W*_*i*(*t*1)_|***A***_*i*(*t*0)_, ***L*_*i*_**) by fitting models regressing *W*_*i*(*t*1)_on ***A***_*i*(*t*0)_and ***L*_*i*_**.

**Step 5**. The decision criteria are used to define treatment 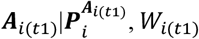. Then refit **Models 1**−**4** to construct 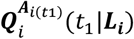 and estimate 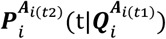. Then estimate *W*_*i*(*t*2)_and determine treatment 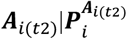. Repeat steps 1-4 to estimate 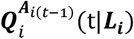 and 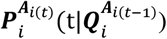 at each t until the end *t*_*e*_.

For participant *i*, the counterfactual SSLY in substate *k* under the dynamic treatment regime 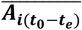 can be estimated as

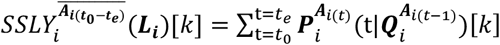

For participant *i*, the counterfactual MALY can be estimated as

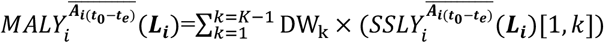

For participant *i*, the counterfactual disease path in substate *k* under the dynamic treatment regime 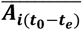 can be estimated as

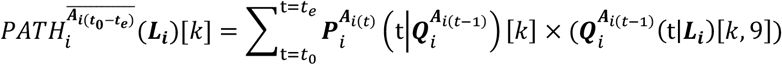

The counterfactual SSLY in the whole population can be estimated as

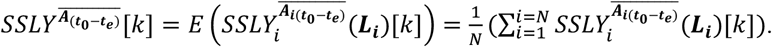

The counterfactual MALY in the whole population can be estimated as

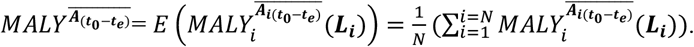

The counterfactual disease path in the whole population can be estimated as

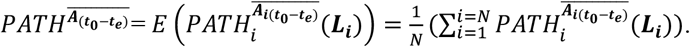

### 2.6. Compare counterfactual MALY across treatment regimes

The difference in MALY between two treatment regimes can be estimated as

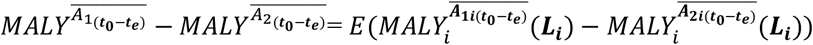

The 95% confidence interval (CI) can be estimated using bootstrap. We randomly draw samples with replacement to create a new sample that are the same size as the original population. The counterfactual MALY for each treatment regime and the difference in MALY between two regimes can be estimated from each bootstrap sample. Statistically, the resulting parameter estimates across all bootstrap samples represent the empirical distribution and can be summarized with median values and a 95% confidence interval (CI) defined by 2.5th to 97.5th percentiles of the empirical distribution.

## 3. APPLICATION

We applied our method to evaluate the cardiovascular benefits of smoking cessation across the course of heart disease in the Atherosclerosis Risk in Communities (ARIC) study.

### 3.1 Study population

We used data from the ARIC study.^14,15^ The enrolled participants underwent a phone interview and clinic visit at baseline in 1987 and were followed up by telephone calls and re-examinations until 2019. Participants were contacted periodically by phone and interviewed about interim hospital admissions, cardiovascular outpatient diagnoses, and deaths. We obtained ARIC data through Biologic Specimen and Data Repository Information Coordinating Center (BioLINCC), an open repository managed by the National Heart, Lung, and Blood Institute (NHLBI). The study was deemed exempt by the Institutional Review Board (IRB) of the University of North Carolina (UNC) at Chapel Hill.

### 3.2. Assessment of exposure, outcome, and covariates

1) Exposure. Smoking status (current smoker, past smoker, never smoker) was assessed using questionnaires at each visit. 2) Variables used to define disease states. Low density lipoprotein cholesterol (LDL-c), high density lipoprotein cholesterol (HDL-c), triglycerides, and blood glucose were assessed in laboratories at each visit. Blood pressure was measured using electronic monitor by medical professionals at each visit. Participants who reported CVD-related events were asked to provide medical records that were reviewed by physicians.^15^ Deaths were identified from systematic searches of vital records in states and the National Death Index, supplemented by reports from family members and postal authorities.^16^ 3) Covariates. Age, sex, race, education level, and family history of heart disease were measured using questionnaires at baseline. Body mass index (BMI) was measured on weight scales by medical professionals.

### 3.3. Definition of the course of heart disease

We modeled the course of heart disease in five states: Healthy, at metabolic risk, coronary heart disease (CHD), heart failure, and mortality. The at-metabolic-risk state was defined as the development of hypertension, hyperlipidemia, or diabetes. Hypertension was defined as blood pressure ≥140/90 mmHg or a history of hypertension or use of blood pressure medications.^17^ Hyperlipidemia included primary hypertriglyceridemia (≥175 mg/dL) or primary hypercholesterolemia (LDL-c 160–189 mg/dL, and/or non-HDL-c 190– 219 mg/dL).^18^ Diabetes was defined as fasting glucose ≥7.7mmol/L.^19^

### 3.4. Regimes of smoking status

We compared four smoking regimes as below. Regimes 1-3 were static regimes which predefined smoking status before intervention. Regime 4 was a dynamic regime defining smoking status by a person’s characteristics that can change over time.

Regime 1: Being a smoker in all disease states.

Regime 2: Being a smoker in healthy and at-metabolic-risk states and stopping smoking if the person develops CHD and heart failure.

Regime 3: No smoking in all disease states.

Regime 4: Being a smoker at the start of intervention and stopping smoking if age >65y.

### 3.5. Statistical analysis

We applied our method to examine the effects of smoking regimes on the course of heart disease. The modeled fitted can be found in the **supplemental materials**. In brief, the transition rates between substates were estimated using cause-specific Cox models, adjusting for age (continuous), sex (male, female), race (White, Black), education (basic, intermediate, advanced), family history of heart disease (yes, no), and BMI (<25, 25-30, ≥30 kg/m^2^). For each treatment regime, the age-specific transition rates and state occupation probabilities were estimated, which were used to synthesize MALY, SSLY, and disease path. The most beneficial regime was identified as the one associated with the highest MALY.

### 3.6. Results

Figure 1. shows the associations of smoking status with rates of transitioning to the next states. Starting from the healthy state, being a current smoker was associated with rate ratios of 0.84 (95% CI: 0.75, 0.95), 4.08 (95% CI: 2.34, 7.12), 2.49 (95% CI: 1.56, 3.96), and 2.65 (95% CI: 2.06, 3.42) of transitioning to at-metabolic-risk, CHD, heart failure, and mortality states, respectively. Similarly, we found strong positive associations of smoking with transitioning to the next states starting from at-metabolic-risk, CHD, and heart failure states. This shows that smoking cessation has health benefits at every state over the course of heart disease.

The cardiovascular benefits of smoking cessation regimes are presented in **Table 1**. Compared to Regime 1 “being a smoker in all disease states”, the MALY was 0.53 (95% CI: 0.21, 0.96) for Regime 2 “being a smoker in healthy and at-metabolic-risk states and stopping smoking in CHD and heart failure states”, 6.10 (95% CI: 4.88, 7.19) for Regime 3 “no smoking in all disease states”, and 4.34 (95% CI: 3.02, 5.47) years higher for Regime 4 “being a smoker at the start of intervention and stopping smoking if age >65y”. Thus, the optimal smoking cessation regime was Regime 3, “no smoking in all disease states”, showing that the primary prevention of heart disease by quitting smoking from the very beginning generated the most cardiovascular benefits.

**Table 1.**
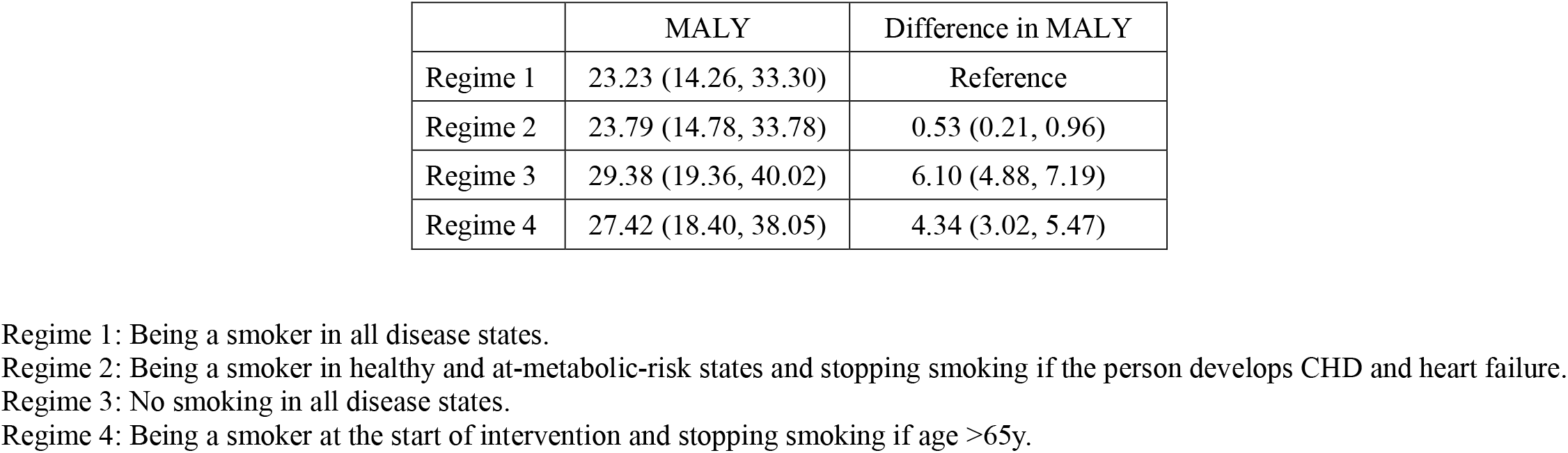
Difference in multimorbidity-adjusted life years (MALY) across smoking regimes in the Atherosclerosis Risk in Communities (ARIC) Study starting from 1987 to 2019 (n=15027).

To investigate the mechanisms by which smoking cessation regimes affect disease progression, we estimated SSLY and disease path under each treatment regime. As shown in **Table 2** and **Figure 3,** compared to Regime 1, the SSLY of the optimal regime, Regime 3, was higher in all states, particularly in the healthy and at-metabolite-risk states. The SSLY was higher for CHD- and heart failure-related substates for Regime 2 in comparation to Regime 1, showing that quitting smoking after the development of CHD or heart failure (which is secondary prevention) elongates life expectancy. The SSLY was higher in all substates under Regime 4, suggesting that quitting smoking at a certain age generates health benefits in all disease states. As shown in **Table 3**, compared to other regimes, Regime 3 had a lower proportion of participants transitioned through the path “Healthy → mortality”, indicating that smoking cessation lowered the risks of mortality through non-cardiometabolic paths such as lung cancer. Correspondingly, Regime 3 had a higher proportion of participants transitioned through the path “Healthy → at metabolic risk → mortality”.

**Table 2.** Estimated substate-specific life years (SSLY) by smoking regimes in the Atherosclerosis Risk in Communities (ARIC) Study starting from 1987 to 2019 (n=15027).

| Disease substates | Life Year (95% CI) |  |  |  |
| --- | --- | --- | --- | --- |
|  | Regime 1 | Regime 2 | Regime 3 | Regime 4 |
| Healthy (S<sub>0</sub>) | 12.47 (7.90, 17.45) | 12.47 (7.90, 17.45) | 13.72 (9.18, 18.20) | 13.88 (9.21, 18.74) |
| At metabolic risk (S<sub>1</sub>) | 16.78 (4.99, 29.38) | 16.79 (5.00, 29.39) | 22.83 (8.54, 36.16) | 20.86 (7.54, 33.82) |
| CHD without metabolic risk factors (S<sub>2\|0</sub>) | 0.39 (0.07, 1.85) | 0.44 (0.08, 2.11) | 0.16 (0.03, 0.84) | 0.32 (0.06, 1.59) |
| CHD with metabolic risk factors (S<sub>2\|0_1</sub>) | 1.22 (0.37, 3.25) | 1.39 (0.42, 3.67) | 1.14 (0.41, 2.92) | 1.23 (0.39, 3.31) |
| Heart failure with no metabolic risk factors or CHD (S<sub>3\|0</sub>) | 0.36 (0.14, 0.90) | 0.44 (0.17, 1.08) | 0.39 (0.15, 1.03) | 0.44 (0.17, 1.09) |
| Heart failure only with metabolic risk factors (S<sub>3\|0_1</sub>) | 1.48 (0.49, 2.62) | 1.78 (0.60, 3.16) | 1.82 (0.73, 3.26) | 1.74 (0.65, 3.15) |
| Heart failure only with CHD (S<sub>3\|0_2</sub>) | 0.12 (0.02, 0.59) | 0.13 (0.03, 0.67) | 0.05 (0.01, 0.28) | 0.11 (0.02, 0.54) |
| Heart failure with metabolic risk factors and CHD (S<sub>3\|0_1_2</sub>) | 0.33 (0.07, 0.87) | 0.38 (0.08, 0.99) | 0.32 (0.08, 0.86) | 0.35 (0.08, 0.95) |

**Table 3.**
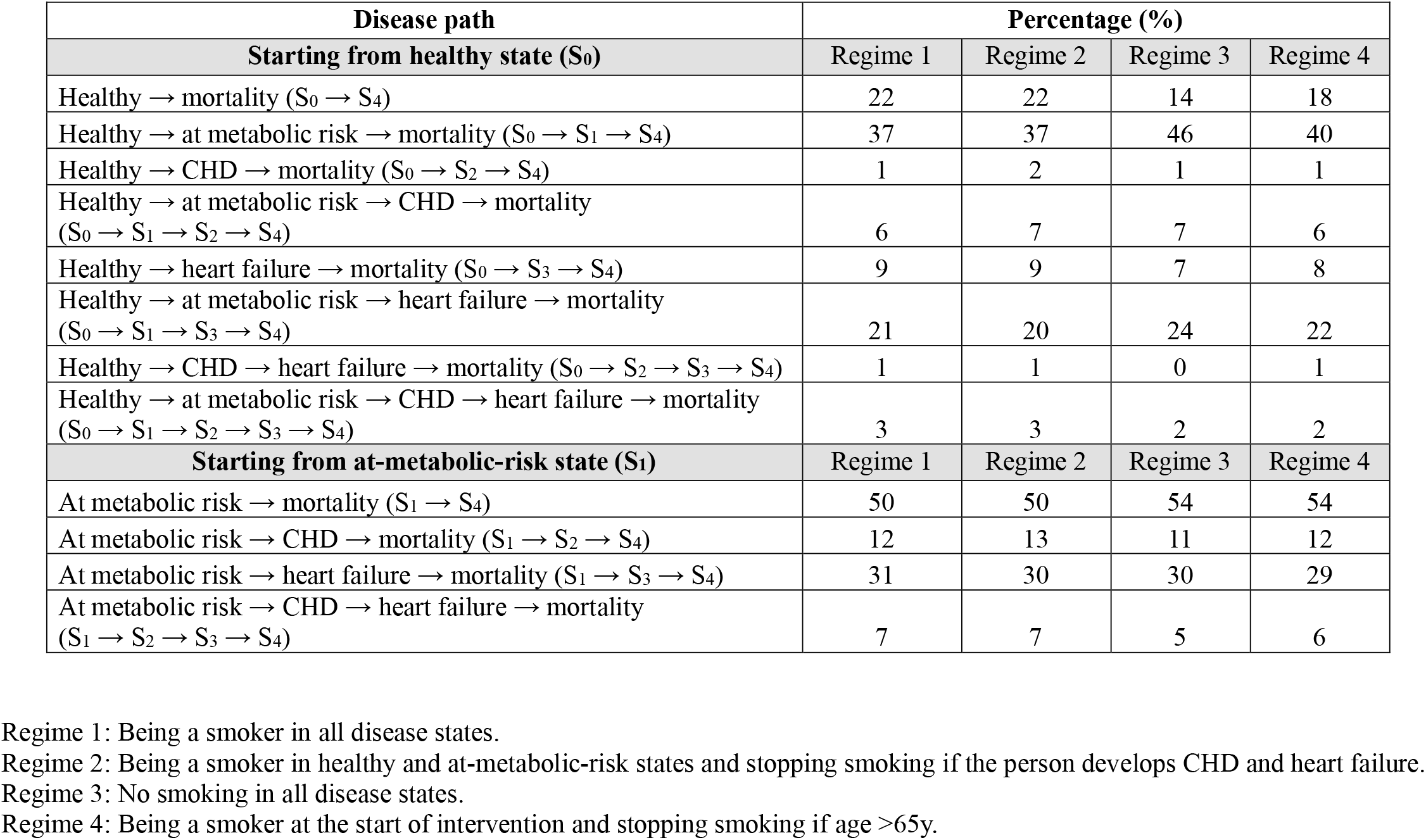
Estimated disease path by smoking regimes in the Atherosclerosis Risk in Communities (ARIC) Study starting from 1987 to 2019 (n=15027).

**Figure 3.**
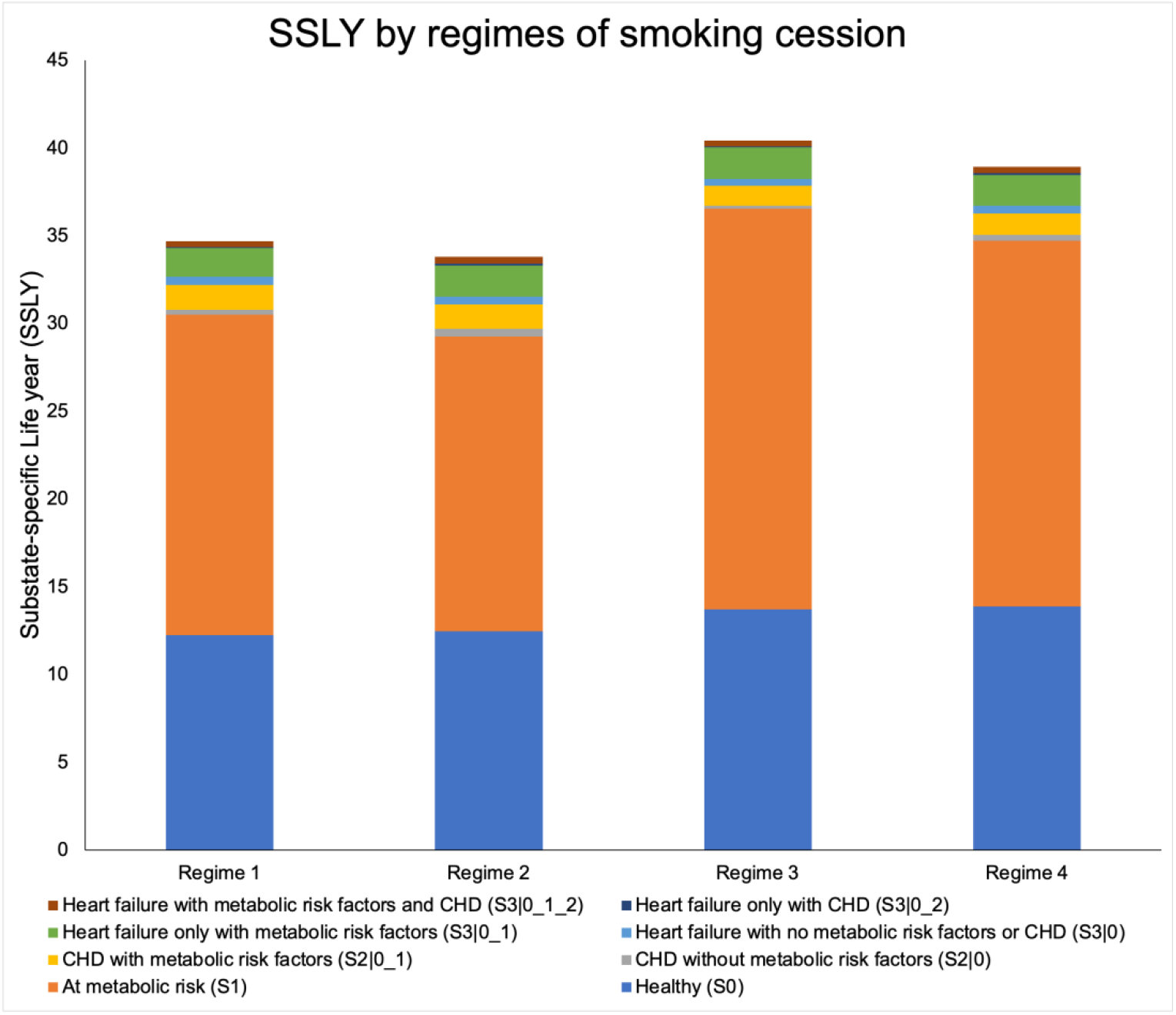
Estimated substate-specific life years (SSLY) by smoking regimes in the Atherosclerosis Risk in Communities (ARIC) Study starting from 1987 to 2019 (n=15027). Regime 1: Being a smoker in all disease states. Regime 2: Being a smoker in healthy and at-metabolic-risk states and stopping smoking if the person develops CHD and heart failure. Regime 3: No smoking in all disease states. Regime 4: Being a smoker at the start of intervention and stopping smoking if age >65y.

## 4. DISCUSSION

The method is developed based on a Discrete-time Split-state Framework for multi-state modeling.^6^ The Framework splits disease states into substates by conditioning on past history and the newly created substates have two unique features. First, memoryless. By conditioning on past history, the substates are independent of past states and satisfy the Markov assumption. Regardless of whether the original process is Markovian, the newly created substates exhibit Markov properties so that the Aalen-Johansen estimator can be used to estimate state occupation probability based on the transition rates. Second, memorable. While the substates satisfy the Markov assumption, they contain information of past history. Being in a substate indicates multimorbidity status and previous disease path. The “memoryless” and “memorable” features allow us to synthesize the substate- and time-specific transition rates into summary measures (i.e., SSLY, MALY, and disease path) to characterize the entire disease course. In this paper, based on this framework, we propose a multi-state causal framework to identify the optimal treatment regime that generates the most benefits as indicated by the maximum MALY. Our method also illustrates the mechanisms of treatment regimes on disease progression as indicated by SSLY and disease path. For public health significance, our method has the potential to advance the precision prevention of chronic diseases by identifying the optimal treatment at “each” (sub)state for “each” person that maximizes the “precise” benefits over disease course.

From a methodology perspective, our method is a causal inference approach that evaluates the counterfactual effect of a treatment regime on the disease course, assuming all participants receive the treatment. Existing methods that examine the effects of treatment regimes on disease risk include the parametric g-formula and the marginal structure model (MSM).^1,2^ Both methods control for time-varying confounding by conditioning on past confounders under treatment and estimate conditional survival probabilities using the Kaplan–Meier estimator. The counterfactual effect in the entire population is obtained through standardization (for g-formula) or inverse probability weighting (for MSM). However, these methods reply on the risk of a single endpoint to make decisions, and omitting the disease course may lead to misidentifying the optimal treatment, particularly when a treatment has long-term effects on multiple disease states. Our method takes the entire disease course into account and can be considered as an extension of g-formula in the multi-state setting. In addition to causal inference methods, Sequential Multiple Assignment Randomized Trials (SMARTs) has been used to evaluate the causal effect of treatment regimes on an outcome.^20^ In a SMART trial, participants are randomized multiple times to formulate decision rules to build optimal adaptive treatment strategies. However, a sequential trial is impractical for chronic diseases in most cases, as an extreme long follow-up period is needed for the multi-states to develop. By evaluating the counterfactual effect of a treatment regime, our method emulates a hypothetical sequential trial that would be difficult to conduct in practice.

From a public health perspective, our method has the potential to advance the precision prevention of chronic diseases by identifying the optimal treatment for each person at each (sub)state that maximizes the precise benefits over the disease course. First, our method allows for the identification of optimal (sub)state-specific treatments. Traditionally, chronic disease prevention includes primary and secondary prevention: primary prevention aims to prevent disease incidence, and secondary prevention aims to prevent disease complications.^21^ Given the complexity of chronic disease progression, prevention strategies may need to vary further based on disease states and past history. Our method enables the specification of substate-specific strategies for precision prevention. Second, our method allows for dynamic interventions tailored to a person. Traditionally, public health prevention strategies are applied to the general population, with individual variability not considered. In this paper, our dynamic regime accounts for individual heterogeneity in assigning treatments, as our method allows to make treatment decisions based on a person’s disease states and characteristics over time, such as responses to treatment. Our method enables the identification of personalized prevention strategies that are most beneficial across the entire disease course.^2^ Third, because MALY takes multimorbidity and quality of life into account, our method provides more precise estimation of prevention effects, which can be used for cost-effectiveness analysis and inform decision-making. Furthermore, the estimated SSLY and disease path illustrate the mechanisms of treatment on disease progression. As shown in the application section, never smokers (Regime 3) significantly increased MALY primarily by elongating the at-metabolic-risk state compared to other Regimes, demonstrating that earlier smoking cessation yields better prevention results. Moreover, compared to other regimes, Regime 3 had a lower proportion of participants transitioned through the path “Healthy → mortality”, indicating that smoking cessation lowered the risks of mortality through non-cardiometabolic paths such as lung cancer.

We acknowledge that our method relies on the assumptions that the effect of treatments on transition rates are instantaneous, that the side effects of treatments are instantaneous, and that confounders are constant over follow-up. Although the disease substates created using our framework contain information about past history, they do not track past history to inform time spent in each past substates. If treatments had cumulative effects, we would need to further identify the time spent in each past substate to estimate the cumulative treatment effect, as treatments can differ by disease substate. This, however, is beyond the scope of this paper (There are two scenarios where our proposed method can be applied to treatments with cumulative effects: one is a static treatment regime with treatments that are the same for all disease states, and the other is a two-state transition). Similarly, for time-varying confounders that can be influenced by treatment or disease states, we would need to track a person’s past history to determine the time spent in each substate to model these confounders over time. For future research, it is of high interest to investigate how time-varying confounders influence the causal effects of treatment regimes on disease progression. Moreover, it is warranted to estimate causal MALY and SSLY while allowing treatment regimes to have cumulative effects on disease progression.

In summary, we have developed a multi-state causal framework to estimate the health benefits of static and dynamic treatment regimes over the disease course. Our method can identify the optimal treatment regime that generates the most benefits as indicated by MALY and illustrate the mechanisms of treatment regimes affecting disease progression as indicated by SSLY and disease path. Our method has the potential to improve the precision prevention of chronic diseases.

## Data Availability

All data can be requested through Biologic Specimen and Data Repository Information Coordinating Center (BioLINCC), an open repository managed by the National Heart, Lung, and Blood Institute (NHLBI).

## Reference

1. Robins JM, Hernan MA, Brumback B. Marginal structural models and causal inference in epidemiology. Epidemiology. 2000;11:550–560. doi: 10.1097/00001648-200009000-00011

2. Young JG, Cain LE, Robins JM, O’Reilly EJ, Hernan MA. Comparative effectiveness of dynamic treatment regimes: an application of the parametric g-formula. Stat Biosci. 2011;3:119–143. doi: 10.1007/s12561-011-9040-7

3. Olafiranye O, Zizi F, Brimah P, Jean-Louis G, Makaryus AN, McFarlane S, Ogedegbe G. Management of Hypertension among Patients with Coronary Heart Disease. Int J Hypertens. 2011;2011:653903. doi: 10.4061/2011/653903

4. Viau DM, Sala-Mercado JA, Spranger MD, O’Leary DS, Levy PD. The pathophysiology of hypertensive acute heart failure. Heart. 2015;101:1861–1867. doi: 10.1136/heartjnl-2015-307461

5. Meira-Machado L, de Una-Alvarez J, Cadarso-Suarez C, Andersen PK. Multi-state models for the analysis of time-to-event data. Stat Methods Med Res. 2009;18:195–222. doi: 10.1177/0962280208092301

6. Ding M., Chen H., F.C. L. A discrete-time split-state framework for multi-state modeling with application to describing the course of heart disease. BMC Medical Research Methodology. 2025:25–54.

7. Ding M, Lin FC, Meyer ML. Summary Estimates Derived from a Multi-state Non-Markov Framework to Characterize the Course of Heart Disease. BMC Medical Research Methodology (in revision) medRxiv 2024.

8. Hernán MA, JMR. Causal Inference: What If. Boca Raton: Chapman & Hall/CRC. 2020.

9. Gray RG. A Class of K-Sample Tests for Comparing the Cumulative Incidence of a Competing Risk. The Annals of Statistics. 1988;16:1141–1154

10. Gerds TA, Ohlendorff JS, Blanche P, Mortensen R., Wright M., Tollenaar N., Muschelli J., Mogensen U.B., B. O. Risk Regression Models and Prediction Scores for Survival Analysis with Competing Risks. https://githubcom/tagteam/riskRegression. 2023.

11. Aalen OO, Johansen S. An Empirical Transition Matrix for Non-Homogeneous Markov Chains Based on Censored Observations. Scandinavian Journal of Statistics. 1978;5:141–150.

12. Murray CJ. Quantifying the burden of disease: the technical basis for disability-adjusted life years. Bull World Health Organ. 1994;72:429–445.

13. Mathers CD, Sadana R, Salomon JA, Murray CJ, Lopez AD. Healthy life expectancy in 191 countries, 1999. Lancet. 2001;357:1685–1691. doi: 10.1016/S0140-6736(00)04824-8

14. The Atherosclerosis Risk in Communities (ARIC) Study: design and objectives. The ARIC investigators. Am J Epidemiol. 1989;129:687–702.

15. Rose GA. Cardiovascular survey methods. Geneva Albany, N.Y.: World Health Organization; WHO Publications Centre distributor. 1982.

16. Fung TT, Chiuve SE, McCullough ML, Rexrode KM, Logroscino G, Hu FB. Adherence to a DASH-style diet and risk of coronary heart disease and stroke in women. Archives of internal medicine. 2008;168:713–720. doi: 10.1001/archinte.168.7.713

17. Unger T, Borghi C, Charchar F, Khan NA, Poulter NR, Prabhakaran D, Ramirez A, Schlaich M, Stergiou GS, Tomaszewski M, et al. 2020 International Society of Hypertension Global Hypertension Practice Guidelines. Hypertension. 2020;75:1334–1357. doi: 10.1161/HYPERTENSIONAHA.120.15026

18. Grundy SM, Stone NJ, Bailey AL, Beam C, Birtcher KK, Blumenthal RS, Braun LT, de Ferranti S, Faiella-Tommasino J, Forman DE, et al. 2018 AHA/ACC/AACVPR/AAPA/ABC/ACPM/ADA/AGS/APhA/ASPC/NLA/PCNA Guideline on the Management of Blood Cholesterol: Executive Summary: A Report of the American College of Cardiology/American Heart Association Task Force on Clinical Practice Guidelines. Circulation. 2019;139:e1046–e1081. doi: 10.1161/CIR.0000000000000624

19. American Diabetes A. Diagnosis and classification of diabetes mellitus. Diabetes Care. 2010;33 Suppl 1:S62–69. doi: 10.2337/dc10-S062

20. Murphy SA. An experimental design for the development of adaptive treatment strategies. Stat Med. 2005;24:1455–1481. doi: 10.1002/sim.2022

21. Arnett DK, Blumenthal RS, Albert MA, Buroker AB, Goldberger ZD, Hahn EJ, Himmelfarb CD, Khera A, Lloyd-Jones D, McEvoy JW, et al. 2019 ACC/AHA Guideline on the Primary Prevention of Cardiovascular Disease: A Report of the American College of Cardiology/American Heart Association Task Force on Clinical Practice Guidelines. Circulation. 2019;140:e596–e646. doi: 10.1161/CIR.0000000000000678

