## Supplementary material for "Estimating the Effects of Treatment Regimes over the Course of Chronic Disease: A Multi-state Causal Framework with Baseline Confounding": Supp

**Supplemental material**

**Statistical analysis of the application study**

**1. Data preparation**. We applied the Discrete-Time Split-State Framework to examine the effects of smoking cession on the course of heart disease. S_0_ to S_4_ states referred to healthy, at-metabolic risk, CHD, heart failure, and mortality states, respectively. We defined substates by conditioning on past disease history: CHD without metabolic risk factors (S_2|0_), CHD with metabolic risk factors (S_2|0_1_), heart failure with no metabolic risk factors or CHD (S_3|0_), heart failure only with metabolic risk factors (S_3|0_1_), heart failure only with CHD (S_3|0_2_), and heart failure with metabolic risk factors and CHD (S_3|0_1_2_). The data were divided into four subsets 1-4, starting from healthy, at-risk, CHD, and heart failure states, respectively. Within each subset, the data were changed from a wide to long format by age. Details of data structure transformation can be found in previous publication.[1]

The four regimes can be written as below.

$\bar{A}_{r1}$= $c$($A_{0}$=1, $A_{1}$=1, $A_{2|0}$=1, $A_{2|1\_0}$=1, $A_{3|0}$=1, $A_{3|1\_0}$=1, $A_{3|2\_0}$=1, $A_{3|2\_1\_0}$=1)

$\bar{A}_{r2}$= $c$($A_{0}$=1, $A_{1}$=1, $A_{2|0}$=0, $A_{2|1\_0}$=0, $A_{3|0}$=0, $A_{3|1\_0}$=0, $A_{3|2\_0}$=0, $A_{3|2\_1\_0}$=0)

$\bar{A}_{r3}$= $c$($A_{0}$=0, $A_{1}$=0, $A_{2|0}$=0, $A_{2|1\_0}$=0, $A_{3|0}$=0, $A_{3|1\_0}$=0, $A_{3|2\_0}$=0, $A_{3|2\_1\_0}$=0)

For $\bar{A}_{r4}$, at each time interval in each of the substate, the intervention was 1 if age $\leq$65 years and changed to 0 if age $>$65 years.

**2. Fit cause-specific Cox models between smoking and the rates of transitioning to next states.**

***Model 5*** *in subset 1 starting from the S_0_ state: h_(Sm=1|S0=1)_(τ)=h_0m_(τ)exp(spline(age) + smoking + sex + race/ethnicity + education + income + family history of heart disease + BMI)*

Model 5 estimated transition rates from state 0 to states 1-4, namely, λ_01_(age|$A_{0}$, $\mathbf{L}$), λ_02_(age|$A_{0}$, $\mathbf{L}$), λ_03_(age|$A_{0}$, $\mathbf{L}$), and λ_04_(age|$A_{0}$, $\mathbf{L}$), where $A_{0}$referred to smoking status (never, past, current) at the start of S_0_ state, and $\mathbf{L}$ were baseline covariates including sex, race/ethnicity, education, income, family history of heart disease, and BMI.

***Model 6*** *in subset 2 starting from S_1_ state: h_(Sm=1|S1=1, S0=1)_(τ)=h_0m_(τ)exp(spline(age) + smoking + sex + race/ethnicity + education + income + family history of heart disease + BMI)*

Model 6 estimated transition rates from state 1 to states 2-4, namely, λ_12_(age|$A_{1}$, $\mathbf{L}$), λ_13_(age|$A_{1}$, $\mathbf{L}$), and λ_14_(age|$A_{1}$, $\mathbf{L}$), where $A_{1}$referred to smoking status (never, past, current) at the start of S_1_ state.

***Model 7*** *in subset 3 starting from S_2_ state:*

*h_(Sm=1|S2=1)_(τ)=h_0m_(τ)exp(spline(age) + past history of S_1_ + smoking + sex + race/ethnicity + education + income + family history of heart disease + BMI)*

Model 7 estimated transition rates from substate S_2|0_ to state 3-4, namely, λ_23|0_(age|$A_{2|0}$, $\mathbf{L}$) and λ_24|0_(age|$A_{2|0}$, $\mathbf{L}$), and transition rates from substate S_2|0_1_ to state 3-4, namely λ_23|0_1_(age|$A_{2|0\_1}$, $\mathbf{L}$) and λ_24|0_1_(age|$A_{2|0\_1}$, $\mathbf{L}$). $A_{2|0}$was smoking status at the start of S_2_ for those without a history of S_1_, and $A_{2|0\_1}$ was smoking status at the start of S_2_ for those with a history of S_1_.

***Model 8*** *in subset 4 starting from S_3_ state:*

*h_(Sm=1|S3=1)_(τ)=h_0m_(τ)exp(age + age^2^ + age^3^ + past history of S_0_ and S_1_ + smoking + sex + race/ethnicity + education + income + family history of heart disease + BMI)*

Model 8 estimated transition rates to S_4_ from substates S_3|0_, S_3|0_1_, S_3|0_2_, and S_3|0_1_2_, namely, λ_34|0_(age$|A_{3|0}$, $\mathbf{L}$), λ_34|0_1_(age|$A_{3|0\_1}$, $\mathbf{L}$), λ_34|0_2_(age|$A_{3|0\_2}$, $\mathbf{L}$), and λ_34|0_1_2_(age|$A_{3|0\_1\_2}$, $\mathbf{L}$). $A_{3|0}$, $A_{3|0\_1}$, $A_{3|0\_2}$, and $A_{3|0\_1\_2}$ were smoking status at the start of S_3_.

**3. Static treatment regimes (**$\boldsymbol{A}_{\boldsymbol{r}\boldsymbol{1}}$**,** $\boldsymbol{A}_{\boldsymbol{r}\boldsymbol{2}}$**,** $\boldsymbol{A}_{\boldsymbol{r}\boldsymbol{3}}$**).**

For each static treatment regime, counterfactual transition rate matrix for participant $i$, $\boldsymbol{Q}_{i}^{\bar{A}}(t|\boldsymbol{L}_{\boldsymbol{i}})$, can be constructed and used to estimate $\boldsymbol{P}_{i}^{\bar{A}}(t|\boldsymbol{L}_{\boldsymbol{i}})$ and ${MALY}_{i}^{\bar{A}}(\boldsymbol{L}_{\boldsymbol{i}}$**).**

The SSLY in substate $k$ is estimated as

${SSLY}_{i}^{\boldsymbol{A}}(\boldsymbol{L}_{\boldsymbol{i}})[k]=\sum_{t=t_{0}}^{t=t_{e}} \boldsymbol{P}_{i}^{\boldsymbol{A}}(t|\boldsymbol{L}_{\boldsymbol{i}})\left[ k \right]$, where $k$ ranges from 1 to 8 except the end state death.

MALY can be calculated as a weighted average of SSLY across the $K$ substates

${MALY}_{i}^{\boldsymbol{A}}(\boldsymbol{L}_{\boldsymbol{i}})$=$\sum_{k=1}^{k=K-1} \mathrm{DW}_{k}\times{SSLY}_{i}^{\boldsymbol{A}}(\boldsymbol{L}_{\boldsymbol{i}})[k]$

The counterfactual disease path in substate $k$ is estimated as

${PATH}_{i}^{\mathbf{A}}(\boldsymbol{L}_{\boldsymbol{i}})[k]=\sum_{t=t_{0}}^{t=t_{e}} \boldsymbol{P}_{i}^{\mathbf{A}}\left( t | \boldsymbol{L}_{\boldsymbol{i}} \right)\left[ k \right]\times(\boldsymbol{Q}_{i}^{\mathbf{A}}\left( t | \boldsymbol{L}_{\boldsymbol{i}} \right)\left[ k,9 \right])$

Specially, the disability weights (DW) were derived from the Global Burden of Disease Study 2019.^16^ The DWs were 0.049, 0.072, and 0.074 for at-metabolic-risk (S_1|0_), CHD (S_2|0_), and heart failure (S_3|0_) states, respectively. For states with multimorbidity, a multiplicative approach was used to assign disability weights. For example, the DW for CHD transitioning from healthy and at metabolic risk states was calculated as: $\mathrm{DW}_{2|0\_1}$=$1-\left( 1-0.049)(1-0.072 \right)$.

The marginal SSLY, MALY, and disease path can be estimated as

${SSLY}^{\boldsymbol{A}}[1,k]$= $E\left( {SSLY}_{i}^{\boldsymbol{A}}(\boldsymbol{L}_{\boldsymbol{i}}\boldsymbol{)}[k] \right)=\frac{1}{N}(\sum_{i=1}^{i=N} {SSLY}_{i}^{\boldsymbol{A}}(\boldsymbol{L}_{\boldsymbol{i}}\boldsymbol{)}[k])$.

${MALY}^{\boldsymbol{A}}$= $E\left( {MALY}_{i}^{\boldsymbol{A}}(\boldsymbol{L}_{\boldsymbol{i}}\boldsymbol{)} \right)=\frac{1}{N}(\sum_{i=1}^{i=N} {MALY}_{i}^{\boldsymbol{A}}(\boldsymbol{L}_{\boldsymbol{i}}\boldsymbol{)})$.

${PATH}^{\boldsymbol{A}}$= $E\left( {PATH}_{i}^{\boldsymbol{A}}(\boldsymbol{L}_{\boldsymbol{i}}\boldsymbol{)} \right)=\frac{1}{N}(\sum_{i=1}^{i=N} {PATH}_{i}^{\boldsymbol{A}}(\boldsymbol{L}_{\boldsymbol{i}}\boldsymbol{)})$.

We estimated ${MALY}^{\boldsymbol{A}_{\boldsymbol{r}\boldsymbol{1}}}$, ${SSLY}^{\boldsymbol{A}_{\boldsymbol{r}\boldsymbol{1}}}$, ${PATH}^{\boldsymbol{A}_{\boldsymbol{r}\boldsymbol{1}}}$, ${MALY}^{\boldsymbol{A}_{\boldsymbol{r}\boldsymbol{2}}}$, ${SSLY}^{\boldsymbol{A}_{\boldsymbol{r}\boldsymbol{2}}}$, ${PATH}^{\boldsymbol{A}_{\boldsymbol{r}\boldsymbol{2}}}$, ${MALY}^{\boldsymbol{A}_{\boldsymbol{r}\boldsymbol{3}}}$, ${SSLY}^{\boldsymbol{A}_{\boldsymbol{r}\boldsymbol{3}}}$, and ${PATH}^{\boldsymbol{A}_{\boldsymbol{r}\boldsymbol{3}}}$.

**4. Dynamic regime of statin use (**$\boldsymbol{A}_{\boldsymbol{r}\boldsymbol{4}}$**)**

**Step 1.** At time $t_{0}$, define state occupation probabilities $\boldsymbol{P}_{\boldsymbol{i}}\mathbf{(}t_{0}|\boldsymbol{L}_{\boldsymbol{i}}$**)**. For example, for a person starting from healthy state, $\boldsymbol{P}_{\boldsymbol{i}}\left( t_{0} | \boldsymbol{L}_{\boldsymbol{i}} \right)\boldsymbol{=c(}1,0,0,0,0,0,0,0,0\boldsymbol{)}$.

**Step 2.** Assign smoking status $\boldsymbol{A}_{i\left( t0 \right)}$ according to age at $t_{0}$.

**Step 3.** Estimate counterfactual transition rates $\boldsymbol{Q}_{i}^{\boldsymbol{A}_{i\left( t0 \right)}}(t_{0}|\boldsymbol{L}_{\boldsymbol{i}})$ by fitting **Models 5-8**.

**Step 5.** Estimate state occupation probabilities $\boldsymbol{P}_{i}^{\boldsymbol{A}_{i\left( t1 \right)}}$ at time $t_{1}$.

$\boldsymbol{P}_{i}^{\boldsymbol{A}_{i\left( t1 \right)}}(t|\boldsymbol{Q}_{i}^{\boldsymbol{A}_{i\left( t0 \right)}})$=$\boldsymbol{P}_{\boldsymbol{i}}\mathbf{(}t_{0}|\boldsymbol{L}_{\boldsymbol{i}})(\boldsymbol{I}_{\boldsymbol{9}}+\boldsymbol{Q}_{i}^{\boldsymbol{A}_{i\left( t0 \right)}}(t_{1}|\boldsymbol{L}_{\boldsymbol{i}}))$.

Repeat steps 2-5 until a participant’s death, defined as $\boldsymbol{P}_{i}^{\boldsymbol{A}_{i\left( te \right)}}$[9]>0.95.

For participant $i$, the counterfactual SSLY in substate $k$ under the dynamic treatment regime $\bar{\boldsymbol{A}_{\boldsymbol{i(}\boldsymbol{t}_{\boldsymbol{0}}\boldsymbol{-}\boldsymbol{t}_{\boldsymbol{e}}\boldsymbol{)}}}$can be estimated as

${SSLY}_{i}^{\bar{\boldsymbol{A}_{\boldsymbol{i(}\boldsymbol{t}_{\boldsymbol{0}}\boldsymbol{-}\boldsymbol{t}_{\boldsymbol{e}}\boldsymbol{)}}}}(\boldsymbol{L}_{\boldsymbol{i}})[k]=\sum_{t=t_{0}}^{t=t_{e}} \boldsymbol{P}_{i}^{\boldsymbol{A}_{i(t)}}(t|\boldsymbol{Q}_{i}^{\boldsymbol{A}_{i\left( t-1 \right)}})\left[ k \right]$

For participant $i$, the counterfactual MALY can be estimated as

${MALY}_{i}^{\bar{\boldsymbol{A}_{\boldsymbol{i(}\boldsymbol{t}_{\boldsymbol{0}}\boldsymbol{-}\boldsymbol{t}_{\boldsymbol{e}}\boldsymbol{)}}}}(\boldsymbol{L}_{\boldsymbol{i}})$=$\sum_{k=1}^{k=K-1} \mathrm{DW}_{k}\times({SSLY}_{i}^{\bar{\boldsymbol{A}_{\boldsymbol{i(}\boldsymbol{t}_{\boldsymbol{0}}\boldsymbol{-}\boldsymbol{t}_{\boldsymbol{e}}\boldsymbol{)}}}}(\boldsymbol{L}_{\boldsymbol{i}})[1,k])$

For participant $i$, the counterfactual disease path in substate $k$ under the dynamic treatment regime $\bar{\boldsymbol{A}_{\boldsymbol{i(}\boldsymbol{t}_{\boldsymbol{0}}\boldsymbol{-}\boldsymbol{t}_{\boldsymbol{e}}\boldsymbol{)}}}$can be estimated as

${PATH}_{i}^{\bar{\boldsymbol{A}_{\boldsymbol{i}\left( \boldsymbol{t}_{\boldsymbol{0}}\boldsymbol{-}\boldsymbol{t}_{\boldsymbol{e}} \right)}}}(\boldsymbol{L}_{\boldsymbol{i}})[k]=\sum_{t=t_{0}}^{t=t_{e}} \boldsymbol{P}_{i}^{\boldsymbol{A}_{i\left( t \right)}}\left( t | \boldsymbol{Q}_{i}^{\boldsymbol{A}_{i\left( t-1 \right)}} \right)\left[ k \right]\times(\boldsymbol{Q}_{i}^{\boldsymbol{A}_{i\left( t-1 \right)}}\left( t | \boldsymbol{L}_{\boldsymbol{i}} \right)\left[ k,9 \right])$.

The counterfactual SSLY in the whole population can be estimated as

${SSLY}^{\bar{\boldsymbol{A}_{\boldsymbol{(}\boldsymbol{t}_{\boldsymbol{0}}\boldsymbol{-}\boldsymbol{t}_{\boldsymbol{e}}\boldsymbol{)}}}}[k]=E\left( {SSLY}_{i}^{\bar{\boldsymbol{A}_{\boldsymbol{i(}\boldsymbol{t}_{\boldsymbol{0}}\boldsymbol{-}\boldsymbol{t}_{\boldsymbol{e}}\boldsymbol{)}}}}(\boldsymbol{L}_{\boldsymbol{i}}\boldsymbol{)}[k] \right)=\frac{1}{N}(\sum_{i=1}^{i=N} {SSLY}_{i}^{\bar{\boldsymbol{A}_{\boldsymbol{i(}\boldsymbol{t}_{\boldsymbol{0}}\boldsymbol{-}\boldsymbol{t}_{\boldsymbol{e}}\boldsymbol{)}}}}(\boldsymbol{L}_{\boldsymbol{i}}\boldsymbol{)}[k])$.

The counterfactual MALY in the whole population can be estimated as

${MALY}^{\bar{\boldsymbol{A}_{\boldsymbol{(}\boldsymbol{t}_{\boldsymbol{0}}\boldsymbol{-}\boldsymbol{t}_{\boldsymbol{e}}\boldsymbol{)}}}}$= $E\left( {MALY}_{i}^{\bar{\boldsymbol{A}_{\boldsymbol{i(}\boldsymbol{t}_{\boldsymbol{0}}\boldsymbol{-}\boldsymbol{t}_{\boldsymbol{e}}\boldsymbol{)}}}}(\boldsymbol{L}_{\boldsymbol{i}}\boldsymbol{)} \right)=\frac{1}{N}(\sum_{i=1}^{i=N} {MALY}_{i}^{\bar{\boldsymbol{A}_{\boldsymbol{i(}\boldsymbol{t}_{\boldsymbol{0}}\boldsymbol{-}\boldsymbol{t}_{\boldsymbol{e}}\boldsymbol{)}}}}(\boldsymbol{L}_{\boldsymbol{i}}\boldsymbol{)})$.

The counterfactual disease path in the whole population can be estimated as

${PATH}^{\bar{\boldsymbol{A}_{\boldsymbol{(}\boldsymbol{t}_{\boldsymbol{0}}\boldsymbol{-}\boldsymbol{t}_{\boldsymbol{e}}\boldsymbol{)}}}}$= $E\left( {PATH}_{i}^{\bar{\boldsymbol{A}_{\boldsymbol{i(}\boldsymbol{t}_{\boldsymbol{0}}\boldsymbol{-}\boldsymbol{t}_{\boldsymbol{e}}\boldsymbol{)}}}}(\boldsymbol{L}_{\boldsymbol{i}}\boldsymbol{)} \right)=\frac{1}{N}(\sum_{i=1}^{i=N} {PATH}_{i}^{\bar{\boldsymbol{A}_{\boldsymbol{i(}\boldsymbol{t}_{\boldsymbol{0}}\boldsymbol{-}\boldsymbol{t}_{\boldsymbol{e}}\boldsymbol{)}}}}(\boldsymbol{L}_{\boldsymbol{i}}\boldsymbol{)})$.

We estimated ${MALY}^{{\bar{\boldsymbol{A}}}_{\boldsymbol{r}\boldsymbol{4}}}$, ${SSLY}^{{\bar{\boldsymbol{A}}}_{\boldsymbol{r}\boldsymbol{4}}}$, and ${PATH}^{{\bar{\boldsymbol{A}}}_{\boldsymbol{r}\boldsymbol{4}}}$ in the whole population.

**5. Identify optimal regime of smoking cession.** The counterfactual MALY was compared across $\boldsymbol{A}_{\boldsymbol{r}\boldsymbol{1}}$**,** $\boldsymbol{A}_{\boldsymbol{r}\boldsymbol{2}}$**,** $\boldsymbol{A}_{\boldsymbol{r}\boldsymbol{3}}$**,** and $\boldsymbol{A}_{\boldsymbol{r}\boldsymbol{4}}$, and the optimal regime of smoking cession was defined as the one associated with the highest MALY. We presented the counterfactual SSLY for each regime to illustrate the mechanisms of why one regime is optimal over the others. The 95% confidence interval of the difference in MALY between treatment regimes was estimated using bootstrap. We conducted bootstrap for 100 times, with the same sample size as the original population for each bootstrap.

[1] Ding M., Chen H., F.C. L. A discrete-time split-state framework for multi-state modeling with application to describing the course of heart disease. BMC Medical Research Methodology. 2025:25-54.
